## Supplementary material for "Acceptability of a nurse-led non-pharmacological complex intervention for knee pain: Nurse and patient views and experiences": S1 Supplementary file.Interview guide for participants.pdf

**Semi-structured Interview guide for participants.**

### Experience of treatment for knee pain prior to the study

First of all, I would like to talk about any treatments you may have received for your knee pain before the treatment that the nurse gave you here.

- Can you tell me about what treatments you have previously received for your knee pain, before seeing the nurse?
- How did you find following those treatments?
    - Prompts - easy / difficult.

  Probe why easy / difficult

- How suitable did you find the treatment you received?
    - Please elaborate.
        - Attractive? / met your needs?
- Did you have any concerns about the potential benefits and side-effects of the treatments you received?
    - What benefits / side effects?
    - Length of treatment before the study (painkillers)
    - If yes, what concerns?

### Patients' experience of intervention

I would now like to move on to talk about the treatment sessions you received from the nurse and what you thought about them.

#### Relevance and content

- How useful did you find the sessions with the nurse
    - Probe into what specific advice the patient was given on exercise, weight loss and other adjunctive advice, written advice in booklet and how they found it (usefulness), *one by one*
- What did you find to be most useful whilst receiving treatment from the nurse?
    - Probes – exercise sheets, exercise diary, any other materials
- How useful did you find the exercise sheets?
- Were there any aspects that you did not find useful?
    - Probes – *same as above*
- Was there anything else that you would have liked the nurse to do during the treatment sessions?
    - Probes – gaps in advice, exercise performance (leave more time for patient to answer)

#### Provider and delivery

- How did you feel receiving this treatment from a nurse compared to a physiotherapist or doctor? (Reference to patients past treatment).
- What did you think about the nurses' ability to communicate about how to manage your knee pain compared to the healthcare professional met before?
    - Prompts - was the advice / information easy to understand / clear / difficult to understand?
    - Follow up on communication
    - Prompts on education / weight loss
    - Probe - examples
- How did you find the pace of the sessions?
    - Prompts –too slow/fast, any awkward moments?

### Lifestyle changes and following the advice

I would now like to move onto how you managed to follow the advice given to you by the nurse once the treatment sessions finished.

- How did you find following the advice given by the nurse once you had completed the treatment sessions?
    - Probe – easy/difficult – exercises, weight loss, adjunctive advice
- Can you tell me about what changes you have made to your daily life as a result of taking part in the study?
    - Probe – why changes were made/were not made, ease/difficulty of making these changes
- Have you slowly increased the intensity and frequency of the exercises?
- Have you involved any family members or friends in helping you to manage your knee pain? (social life)

  Probe – what they have helped with, why / why not involved anyone

- Going forward, how do you feel about your ability to manage your knee pain?
    - Now
    - In the long-term.

  Probe – confidence in continuing with exercises / weight loss / using other adjunctive treatments

### Changes to perceptions of osteoarthritis and knee pain after the study

- How has your knowledge of osteoarthritis and management of knee pain changed since receiving treatment from the nurse?

  Prompts– *Core treatments* / Was the information too much? / too less to carry home? Could you take it all, remember and carry on?

  Prompt – *add prompts based upon the nurses training booklet and information they tell the patient in the treatment sessions (e.g. importance of paced exercise)*

### Overall satisfaction / sequence of treatment

- In general, how satisfied are you with the treatment that you received from the nurse? Is there anything else that can be improved? (*number or words*)
- To what extent do you feel the treatment programme has met your needs?

**Is there anything else you would like to say?**
